# Hospital-level work organization drives the spread of SARS-CoV-2 within hospitals: insights from a multi-ward model

**DOI:** 10.1101/2021.09.09.21262609

**Authors:** Ajmal Oodally, Pachka Hammami, Astrid Reilhac, Guillaume Guérineau de Lamérie, Lulla Opatowski, Laura Temime

## Abstract

extensive protective measures, SARS-CoV-2 widely circulates within healthcare facilities, posing a significant risk to both patients and healthcare workers. Several control strategies have been proposed; however, the global efficacy of local measures implemented at the ward level may depend on hospital-level organizational factors. We aimed at better understanding the role of between-ward interactions on nosocomial outbreaks and their control in a multi-ward psychiatric hospital in Western France. We built a stochastic compartmental transmission model of SARS-CoV-2 in the 24-wards hospital, accounting for the various infection states among patients and staff, and between-ward connections resulting from staff sharing. We first evaluated the potential of hospital-wide diffusion of local outbreaks, depending on the ward they started in. We then assessed control strategies, including a screening area upon patient admission, an isolation ward for COVID-19 positive patients and changes in staff schedules to limit between-ward mixing. Much larger and more frequent outbreaks occurred when the index case originated in one of the most connected wards with up to four times more transmissions when compared to the more isolated ones. The number of wards where infection spreads was brought down by up to 53 % after reducing staff sharing. Finally, we found that setting up an isolation ward reduced the number of transmissions by up to 70 %, while adding a screening area before admission seemed ineffective.

**Significance Statement:** Hospital acquired COVID-19 poses a major problem to many countries. Despite extensive protective measures, transmission within hospitals still occurs regularly and threatens those essential to the fight against the pandemic while putting patients at risk. Using a stochastic compartmental model, we simulate the spread of SARS-CoV-2 in a multi-ward hospital, assessing the effect of different scenarios and infection control strategies. The novelty of our method resides in the consideration of staff sharing data to better reflect the field reality. Our results highlight the poor efficiency of implementing a screening area before hospital admission, while the setting up of an isolation ward dedicated to COVID-19 patients and the restriction of healthcare workers movements between wards significantly reduce epidemic spread.

## 1. Introduction

The COVID-19 pandemic, caused by severe acute respiratory syndrome coronavirus 2 (SARS-CoV-2), has placed an unprecedented burden on healthcare services worldwide (1),(2),(3). The crisis that followed incited most countries to go into partial or full lockdowns in an effort to curb the spread of the virus. Hospitals and Long Term Care Facilities (LTCFs) have been hit hard, with the former being on the front line to deal with the epidemic and the latter having to deal with repercussions on often vulnerable patients while not always being sufficiently prepared (4), (5), (6). Despite most countries doing their best to ramp up vaccination efforts in these healthcare institutions and among the healthcare worker (HCW) population, the spread of variants of concern which could evade vaccination is an ongoing issue (7), (8). In this context, within-hospital transmission, henceforth referred to as nosocomial transmission, can drastically impact day-to-day operations while putting both HCWs and vulnerable patients at risk. There has been a staggering number of SARS-CoV-2 outbreaks in LTCFs and hospitals with often devastating consequences for elderly patients and especially those with comorbidities (9). On 31 March 2021, LTCF residents in France and Belgium accounted for 42 % and 57 % of total COVID-19 related deaths respectively according to surveillance data from the European Centre for Disease Prevention and Control. As of 31 August 2020, residents of LTCFs accounted for 40 % of US COVID-19 related fatalities (10). Patients in LTCFs require constant care putting them in close and frequent contact with HCWs who might unknowingly act as vectors of transmission. On the flip side, Taiwan’s impressive response to COVID-19 includes an efficient and crucial role played by hospitals in mitigating the spread of the infection (11). Similarly, in the US where a massive vaccination campaign significantly reduced the death toll in most states, the implementation of rigorous control measures has also proven effective in a large medical center (12). Hence, better comprehension of transmission pathways and prevention strategies can greatly reduce the extent of nosocomial COVID-19.

Mathematical modeling of the aforementioned phenomena serves as a powerful tool to evaluate the relevance of infection control strategies. Despite the work of many studies on the epidemic risk at the national or regional levels (13),(14),(15),(16),(17), (18), (19), studies focusing on modeling SARS-CoV-2 transmission in the healthcare setting are still limited (20), (21), (22), (23) (24), (25), (26), (27), (28), (29). Most of the latter focus on testing strategies as control measures, and more importantly, none account for the organizational structure of the healthcare facility under study. Better understanding of nosocomial COVID-19 transmission and other possible infection control strategies should therefore be thoroughly investigated with the aim to alleviate the burden on our healthcare institutions.

With this goal in mind, we propose a new SARS-CoV-2 hospital transmission model that accounts for multiple interconnected wards. After feeding this model with data from the main hospital site of a French psychiatric hospital, henceforth simply referred to as the hospital, we simulate different scenarios to assess the effectiveness of control measures on the spread of the epidemic within the hospital, notably underlining the importance of the ward connectivity.

## 2. Results

The model describes the healthcare community as a meta-population divided into *W* = 24 wards located in 8 different buildings. HCWs are composed of doctors, nurses, medical interns, caregivers, maintenance staff and administrative staff. Each ward holds patients and HCWs distributed into several epidemiological compartments representing the natural history of SARS-CoV-2 infection in a discrete manner (Fig. 1a, Fig. 1b). HCWs may be shared between different wards; staff sharing data allows to reconstruct a network of wards connected by HCWs’ care activities (Fig. 1c). In simulation results that follow, patients who were symptomatic on admission and those who became symptomatic during their hospital stay, systematically underwent SARS-CoV-2 RT-PCR testing (sensitivity and specificity as indicated in Table S1 of SI) and were transferred to an isolation ward upon a positive test. Symptomatic HCWs were also underwent RT-PCR testing and took sick leave if their test results were positive. The model was parameterised (see Table S1 of SI) based on data collected in the hospital during the study period and on an outbreak that occurred in ward A2 (Fig. 2).

**Fig. 1.**
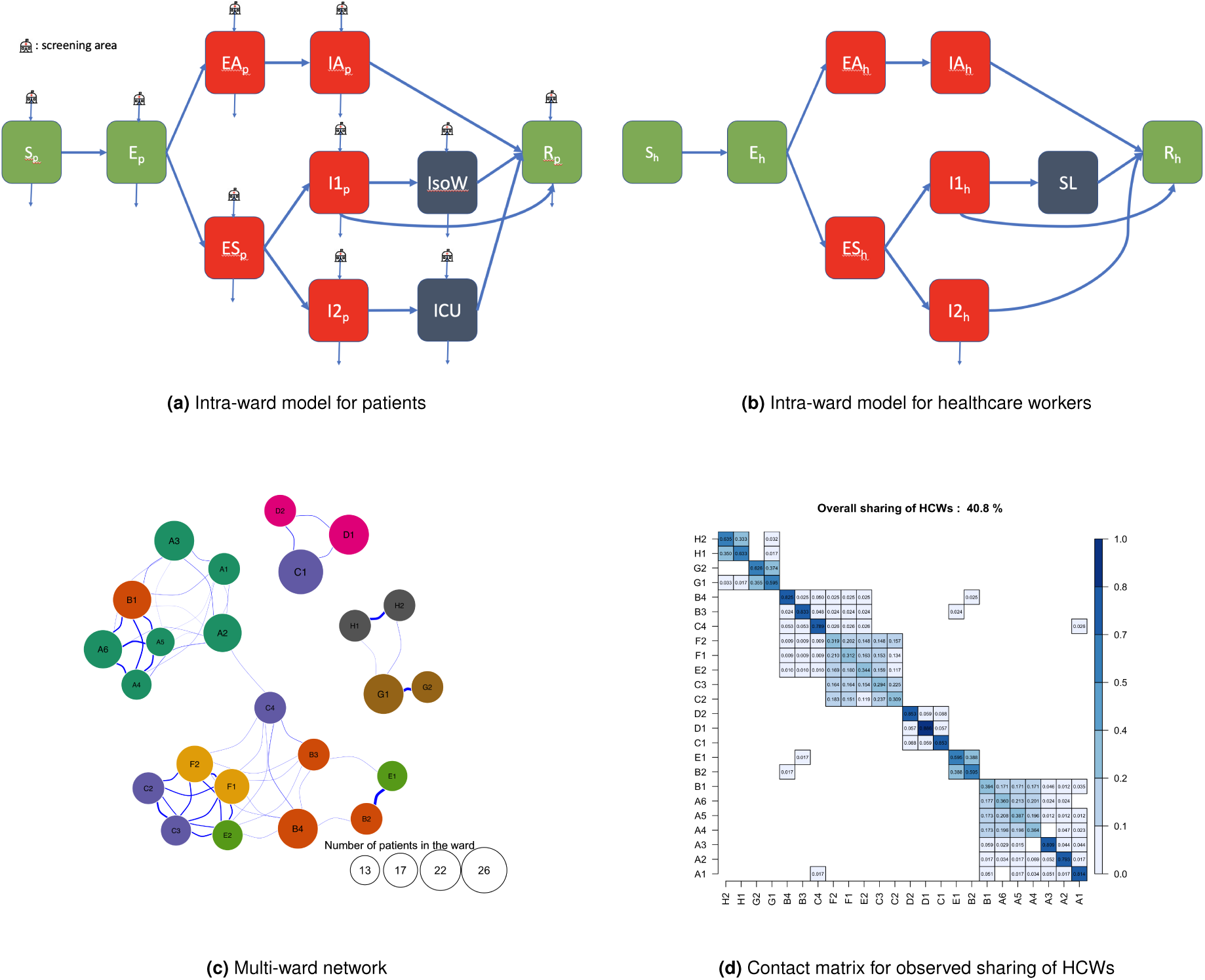
Structure of stochastic multi-ward model. The intra-ward compartmental models for patients and HCWs are represented on the top and middle figure respectively. Red compartments correspond to contagious states while green ones correspond to non-contagious states. Grey compartments represent an intensive care unit (ICU), isolation ward (IsoW) for patients and sick leave (SL) for HCWs. Refer to the material and methods section for a detailed description of each model state. On the bottom figure, the multi-ward model includes all connections resulting from the sharing of HCWs. Wards located in the same building are colored similarly.

**Fig. 2.**
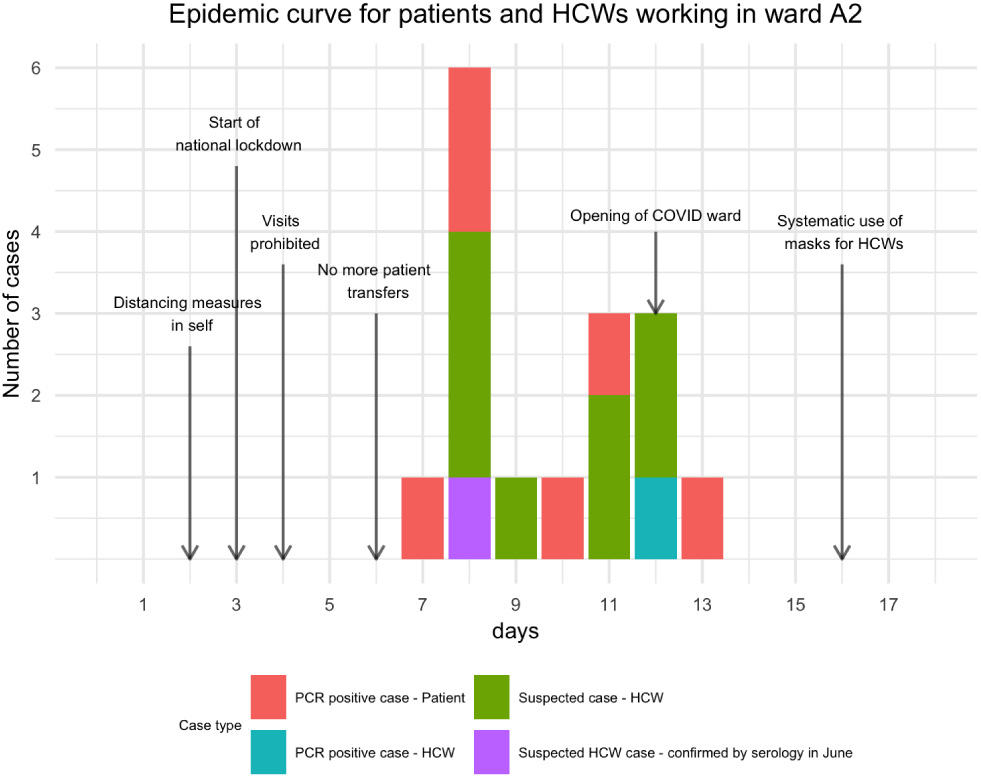
Epidemic curve for ward A2

### A. Ward of origin of index case determines the global risk of outbreak at the hospital level

The connectivity and topology of the ward network may impact the risk and size of an outbreak at the hospital level. Wards were here characterized according to their size and connections to other wards. To better understand and quantify how importations in the different wards can lead to global dissemination risk, we simulated and analyzed the resulting epidemic size following the introduction of a non-contagious incubating patient in one ward at a time. 40 days after the introduction, the median number of nosocomial acquisitions in the entire hospital ranged from two, for an index case introduced in the least connected wards, to sixteen, for an introduction in the most connected one (Fig. 3a). As expected, the number of secondary wards affected was higher when the index ward was more connected (higher degree) (Fig. 3b). In this particular case, three additional interconnections led to one more infected ward on average. Starting from the introduction ward, other wards became infected with probabilities related to their proximity to this initial ward over the network, as illustrated in Fig. 3c-3e.

**Fig. 3.**
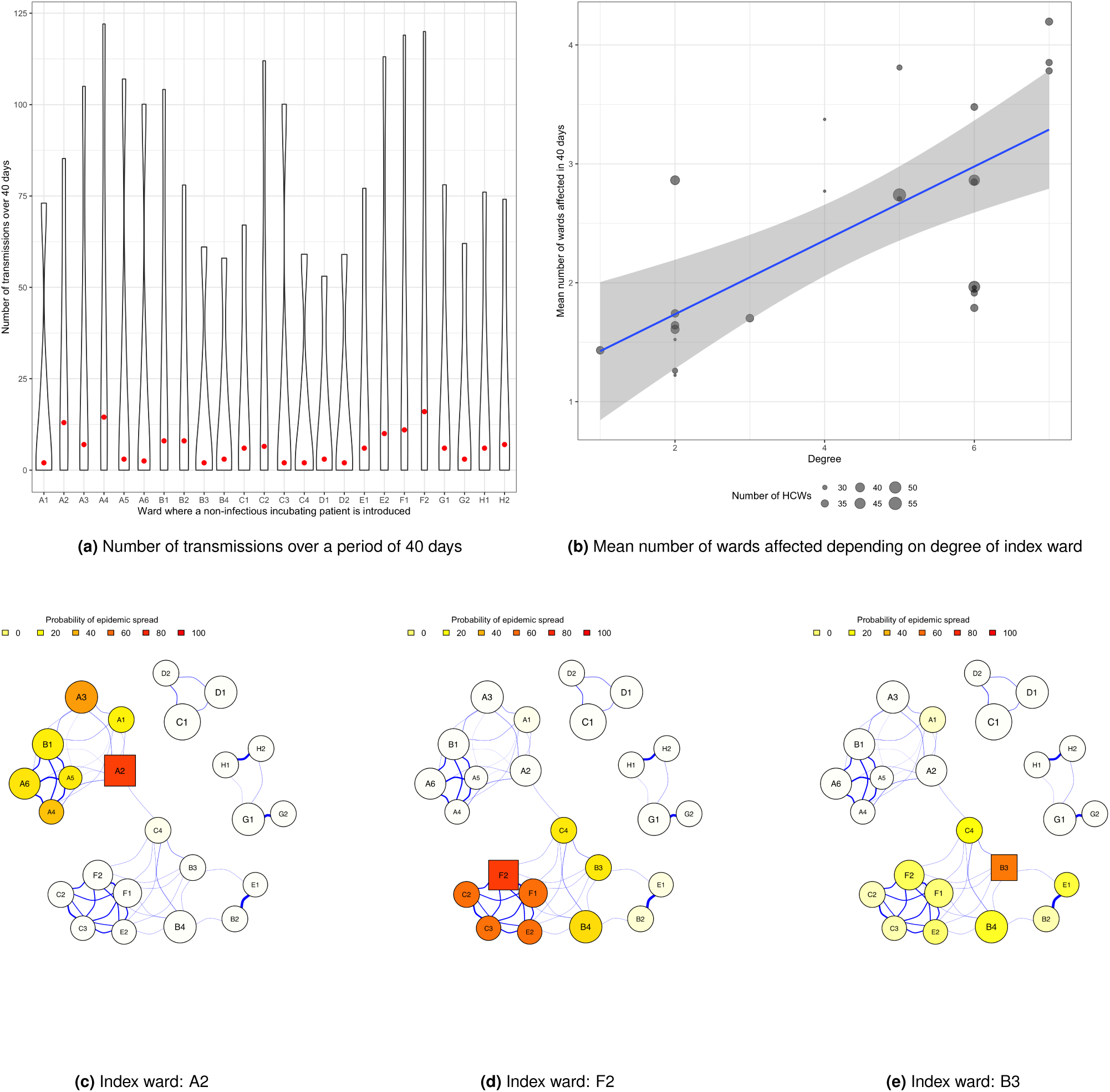
How the index ward determines the global risk at the hospital level. (a) Global number of transmissions in the hospital after 40 days following the introduction of a non-contagious incubating patient in each ward is assessed. The figure shows violin plots based on 500 simulations. Red dots represent the median number of transmissions. (b) Mean number of wards affected by the virus following the introduction of a single case in an index ward, depending on the degree of index ward. (c)-(e) Illustrations of virus spread following introduction in three distinct wards (A2(c), F2(d) and B3(e)).

### B. Reorganizing HCW sharing can impact the epidemic risk

We then explored different work organization scenarios at the hospital level, i.e. different networks, to assess their impact on the epidemic risk. We considered as a baseline scenario (Fig. 1c, 1d) the observed HCW staffing provided by the hospital during the period under investigation. We investigated two hypothetical scenarios: one where between-ward staff sharing was limited by 52.9 % and one where staff sharing was exacerbated by 57.6 %, serving as best-case scenario and worst-case scenario respectively. Limited sharing was modeled by re-assigning HCWs working in multiple wards to fewer wards isolating wards as much as possible; exacerbated sharing was modeled by re-assigning HCWs between wards located in the same building. Both those scenarios implied a different sharing structure of HCWs albeit keeping the same working hours by profession in all wards as in the baseline scenario. Fig. 4a shows the impact of these sharing structures on the propagation of SARS-CoV-2. Particularly useful in cases of low community prevalences, limited staff sharing resulted in up to a 53 % decrease in mean number of wards infected. Oppositely, increasing staff sharing increased up to 44 % the mean number of affected wards. We then assessed the epidemic potential of each ward as point of origin of infection in the three staff organization scenarios. We computed the number of wards where infection spreads and summarized the results based on the degree of the index ward (Fig. 4b). The limited sharing scenario limited the degree of any ward to at most 4, leading to the lowest probability of spread (see Fig. S4, S5 of SI) and outbreaks that never exceed 5 secondary wards, which is not surprising given that most wards were isolated following the reduction in staff sharing (9 out of 24 wards). On the other hand, oversharing of staff frequently led to widespread contamination as illustrated by spates of red in the upper y-values of Fig. 4b.

**Fig. 4.**
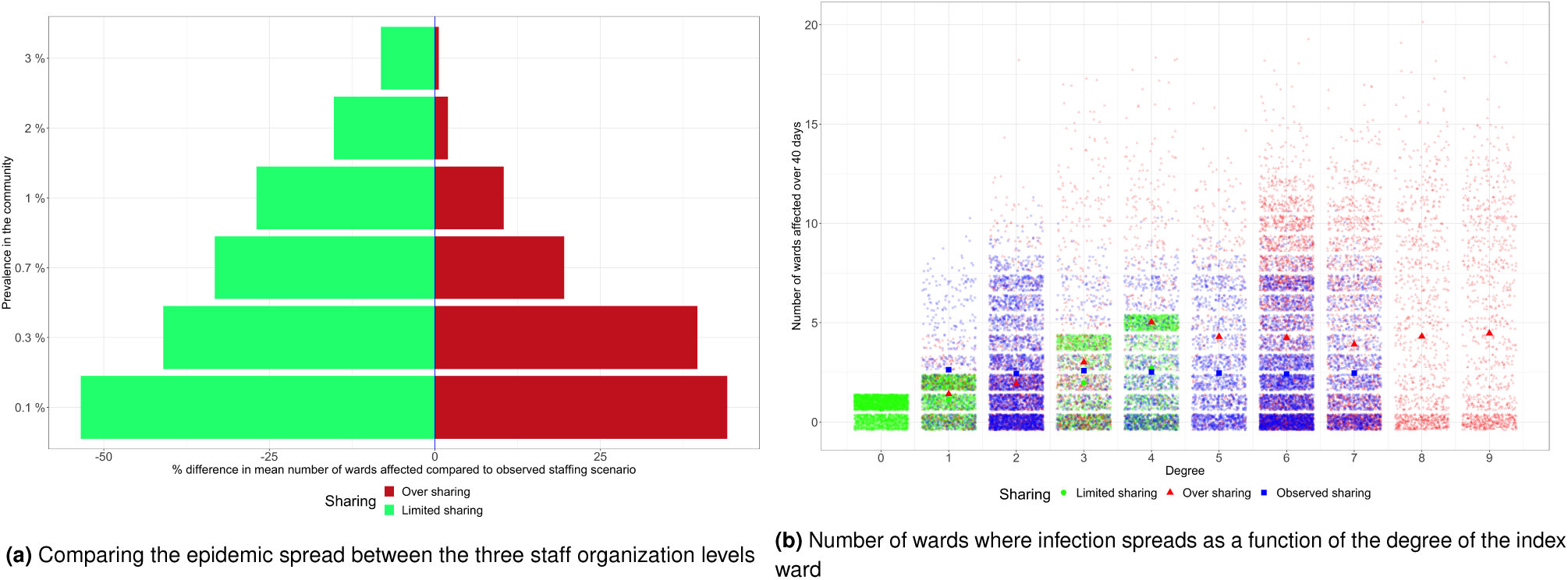
How staff sharing impacts epidemic spread. The figure on the left corresponds to the percentage difference between staff sharing with the observed sharing structure acting as baseline scenario. Y-axis values represent community prevalence values. Results are based on 500 simulations ran over 40 days for each scenario. The figure on the right resumes the results obtained with each ward as index ward and no contamination from the community. Wards with similar degrees and same staff organization level are grouped together. Bold colored shapes represent mean values.

### C. An isolation ward can help reduce outbreak size

We evaluated the efficacy of implementing a COVID-19 isolation ward on preventing the dissemination of SARS-CoV-2 within the hospital. A COVID-19 isolation ward serves as a separate designated space to host detected SARS-CoV-2 positive patients. Symptomatic patients are systematically tested and transferred in case of a positive test. We ran 500 simulations over a 40-day period. Several levels of importation risk were evaluated, assuming different levels of community prevalence (from 0.1% up to 3%, corresponding to a situation close to the epidemic peak during the first pandemic wave). Fig. 5a shows that the presence of a COVID-19 isolation ward consistently led to a lower median number of nosocomial transmissions compared to the reference scenario, absence of a COVID-19 isolation ward. Isolating infected patients substantially brings down the number of transmissions, ranging from a 59 % decrease up to 70 % decrease on average depending on community prevalence. The maximum outbreak potential is also much worse in non-isolation scenarios especially in cases of high community prevalence. Setting up a COVID-19 isolation ward therefore strongly contributes to limiting the dissemination risk in the hospital.

**Fig. 5.**
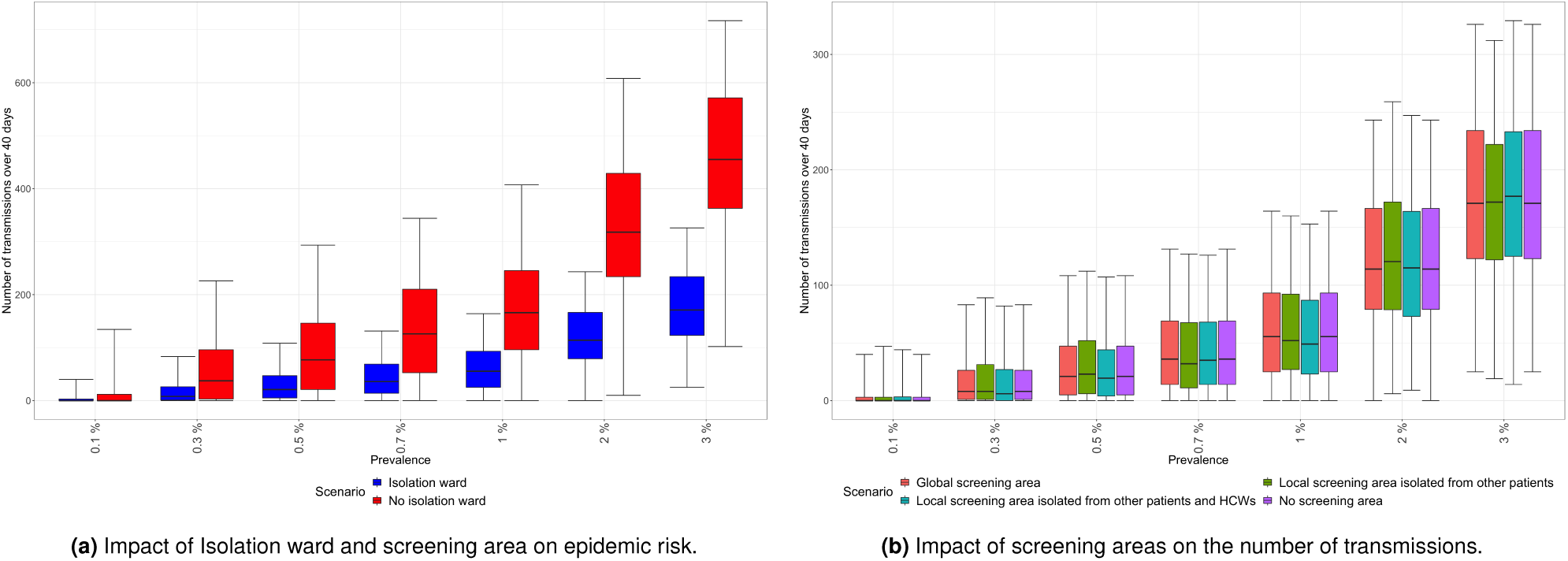
How an isolation ward and screening areas impact epidemic spread. (a) Comparing reference scenarios and a scenario with an isolation ward where positive cases are systematically transferred after 24h on the number of transmissions (y-axis) for different prevalence levels on admission (x-axis) (i.e prevalence in the community) (b) Comparing various screening areas where symptomatic patients are tested, each with a different level of isolation, with a baseline scenario with no screening area on the number of transmissions (y-axis) for different prevalence levels on admission (x-axis). In all scenarios, an isolation ward is implemented.

### D. Screening areas have limited impact on virus transmission

A screening area allows for temporary isolation of newly admitted patients before clinical examination, with RT-PCR tests administered to those presenting symptoms, thereby limiting the admission of positive patients. We assessed the impact of such screening areas and compared them with the reference scenario where symptomatic patients on admission were tested but no screening area was in place. Three types of screening areas were investigated: a global common screening area for the entire hospital, with dedicated staff; a local one for each ward where admitted patients were isolated from other patients from their admission ward; and a local one with isolation from both patients and HCWs of the admission ward. In all these scenarios, a COVID-19 isolation ward was set up and patients who tested positive were assumed to be systematically transferred to the isolation ward within 24 hours. Fig. 5b shows that, for a given community prevalence, the predicted median numbers of nosocomial transmissions after 40 days are quite similar. Insofar as all symptomatic patients were tested and systematically transferred to the isolation ward within 24 hours if they tested positive, screening areas, irrespective of the level of isolation implemented within, showed no impact on the epidemic risk.

## 3. Discussion

In view of the persisting nature of the pandemic and strain on healthcare institutions, all measures likely to limit nosocomial infections should be given serious consideration. In particular, hospital-level work organization plays a significant role in driving the spread of infection and should not be overlooked when designing surveillance and control strategies. The literature on multi-ward models of healthcare-associated infection spread (30) has been on the rise recently, as the necessary contact and transfer data become increasingly available. However, to date, very few studies (23) have taken into account the multi-ward structure of hospitals and LTCFs in their methodology to assess the impact of nosocomial COVID-19 infections. Our study aims at filling this gap and provides an insight in the role of HCW staffing as a major drive of SARS-CoV-2 spread.

In this work, we assessed several infection control strategies aimed at curbing the spread of nosocomial COVID-19 transmissions. Our results indicate that the extent of an outbreak at the hospital level largely depends on the location of the index case of infection. We also showed that the number of connections through HCW sharing was a significant risk factor for widespread contamination. Within 40 days following the introduction of an index case, one additional ward was infected on average following three extra connections. We demonstrated that limiting multi-assignment of HCWs could significantly reduce the risk of epidemic spread throughout the hospital. Finally, while the isolation of infected patients proved to be very effective in curbing the spread of the virus, this was not the case for screening areas.

We found here that HCW rescheduling was an efficient measure to limit nosocomial transmissions and prevent widespread contamination. However, the practical implementation of such a measure, especially if it implies major changes in staff planning, needs to be evaluated. Indeed, while disrupting the routine of already burdened HCWs could prove ill-advised, limiting their multi-assignments could substantially reduce the risk of large scale infection thus contributing to a safer work environment for them as well as patients. A previous study done in the context of healthcare-acquired infections in general came to a similar conclusion (31).

While isolation of identified cases in a dedicated COVID-19 ward was found to be effective, implementation of screening areas focused on testing of symptomatic patients only were found to be ineffective as long as newly symptomatic patients were tested and isolated within 24 hours of admission. Previous studies also point out the effectiveness of an isolation ward and limited impact of measures analogous to a screening area (23), (32).

In order to keep the model as generic and as simple as possible in a context of limited data, we made several assumptions and limitations that should be highlighted. First, we assumed homogeneous mixing within the HCW and patient populations. Regarding HCWs, the risks of exposure are most probably profession-dependent. Similarly, small clusters of contacts may exist within the patient population. The homogeneous mixing assumption may have led us to overestimate the risk of epidemic spread. Second, we estimated transmission rates using data from an outbreak observed in a specific ward and those estimates were then used to characterize all other wards. In doing so, we assumed the same contact patterns n all wards. Third, given that visits were strictly prohibited during the first pandemic wave, we did not account for contamination of patients and HCWs by visitors. In further analyses, this assumption should be relaxed to avoid underestimating the epidemic risk. Also, it would be of interest to account for vaccination in both patient and HCW populations in the model. Vaccine roll-out in LTCFs and hospitals surely plays an important role in further mitigating the spread of infection. Moreover, testing strategies based on network structure could be designed so as to make better use of testing resources in the hospital setting. Following the end of the first lockdown on 11 May 2020, the hospital has been implementing contact tracing to break chains of transmission. Resulting contact data could be used in the model as an infection control measure to limit widespread contamination in case of outbreak. Lastly, a more in-depth and dynamic analysis of the network and its drivers could improve the predictive capacities of our model. Tools such as exponential random graph models (33) or dynamic network analysis taking into account the temporality of contact data could serve such objectives.

We proposed a multi-ward stochastic model at the hospital level to simulate virus transmission and to assess infection control measures with the aim of mitigating the nosocomial spread of SARS-CoV-2. The model serves as a very helpful tool in anticipating the impact of measures to be implemented and therefore contributes in informing decision-makers. While we fed the model with data from a particular healthcare institution, the model remains generic and could be easily implemented with data from other hospitals and LTCFs as input, given that specific data on staff scheduling are available. For instance, other healthcare institutions with different network structures might see better or worse outcomes following reorganization of HCW staffing as compared to the results presented in this paper.

## Materials and Methods

### Epidemiological model description

We build a stochastic compartmental model of SARS-CoV-2 transmission within a multi-ward hospital population under the assumption of homogeneous mixing of populations. Individuals are distributed across compartments according to their infectious status and ward localization. Patients can be: susceptible (Sp), non-contagious incubating (Ep), contagious incubating before asymptomatic condition (EAp), contagious incubating before symptomatic condition (ESp), contagious with asymptomatic condition (IAp), contagious with mild symptoms (I1p), contagious with severe symptoms (I2p) and recovered (Rp). When patients develop severe symptoms, they can be transferred in an intensive care unit (ICU). A ward designated to isolate detected sick patients (IsoW) is also implemented. Finally, to model screening areas aiming at clinical examination and/or virological testing before admission in a ward, 7 other compartments (SASp, SAEp, SAEAp, SAESp, SAIAp, SAI1p, SAI2p, SARp) were incorporated to the model.

Similarly, HCWs can be: susceptible (Sh), non-contagious incubating (Eh), contagious incubating before asymptomatic condition (EAh), contagious incubating before symptomatic condition (ESh), contagious with asymptomatic condition (IAh), contagious with mild symptoms (I1h), contagious with severe symptoms (I2h) and recovered (Rh). HCWs with mild symptoms have a probability *p*_SL_ to take sick leave (SL) while HCWs with severe symptoms leave the model and recover with probability 1 - *p*_*D*_. In case of death, they are removed from the model.

Individuals move from one compartment to another following stochastic transitions computed on the basis of a Gillespie algorithm (34). Susceptible individuals are infected through at risk contacts with contagious individuals (patients or HCWs) from the same ward or connected wards via shared HCWs based on staffing schedules. Each ward was independently modeled and wards were connected through a meta-population system. We assumed that wards were only connected by HCWs through multi-assignments or cover. Patient transfers were excluded in the present analysis. Patients are therefore assumed to only be in contact with other patients and HCWs working in the ward they belong to. The data was collected during the first wave of the pandemic which occurred in March and health practitioners at the hospital confirmed that patient transfers and visits were stopped during that time.

### Model parameters

#### Statistical inference

The model was parametrized, when available, directly from data compiled from the hospital database. Parameter values for which no information was available were fixed from the literature. We refer to Table 1 of SI for an exhaustive list of parameter values and their sources. Transmission rates were estimated to reproduce observed data of suspected or confirmed COVID-19 cases collected during an outbreak that occurred in ward A2 (Fig. 2). The outbreak occurred during the first wave of the pandemic, affecting 6 patients and 10 HCWs. At that time, testing policy only targeted symptomatic patients, who were systematically tested, while testing of HCW was much more complicated for administrative reasons and was not generalized.

A single ward model was fitted to the outbreak data of ward A2 using the pomp package (35) and iterative filtering method (36). The observation model was the cumulative symptomatic infections in the patient and HCW populations, assuming a Poisson measurement model. Parameters were estimated by maximum likelihood, using particle filtering to compute a robust estimate of the likelihood and iterated filtering to maximize it over unknown parameters. Estimations computed from synthetic data generated by the model defined with four transmission rates did not result in the recovery of known parameter values. Those parameters were not identifiable in the model. Consequently, a single transmission rate *β* was estimated with proportionality constraints on transmission rates *β*_HP_, *β*_PP_, *β*_PH_ and *β*_HH_. Multiple combinations of parameters were compared and the set that maximized the likelihood was retained. Besides the transmission rate *β*, we also estimated the number of initial patients (*Ep*_0_) and HCWs (*Eh*_0_) in non-contagious incubation. The date of first infection (*t*_*init*_) was not estimated but fixed to values ranging from 1 to 20 days prior to the first detected case. For each aforementioned value of *t*_*init*_, we estimated parameters *β, Ep*_0_ and *Eh*_0_. The parameter estimates that maximized the likelihood were chosen as best estimates. The model was first tested on simulated data, generated to resemble the outbreak data, to evaluate the capacity to recover known parameter values. We refer to SI for a detailed description of the simulation study and estimation procedure.

## Data Availability

Code and data available upon reasonable request

## ACKNOWLEDGMENTS

The work was supported directly by internal resources from the French National Institute for Health and Medical Research, the Institut Pasteur, the Conservatoire National des Arts et Métiers, and the University of Versailles-Saint-Quentinen-Yvelines/University of Paris-Saclay. This study received funding through the MODCOV project from the Fondation de France grant 106059 as part of the alliance framework “Tous unis contre le virus”, the Université Paris-Saclay (AAP Covid-19 2020) and the French government through its National Research Agency project SPHINX-17-CE36-0008-01. The authors would like to acknowledge the help of the EMEA-MESuRS working group on the nosocomial modeling of SARS-CoV-2 (Audrey Duval, Kévin Jean, Sofía Jijón, Ajmal Oodally, Lulla Opatowski, George Shirreff, David RM Smith, Laura Temime).

## Supplementary Information

The deterministic version of the transmission model can be illustrated using differential equations which describe the dynamics of each compartment (see Table S3 for a description of each compartment) in each time step. Equations in the absence of a screening area are given below.

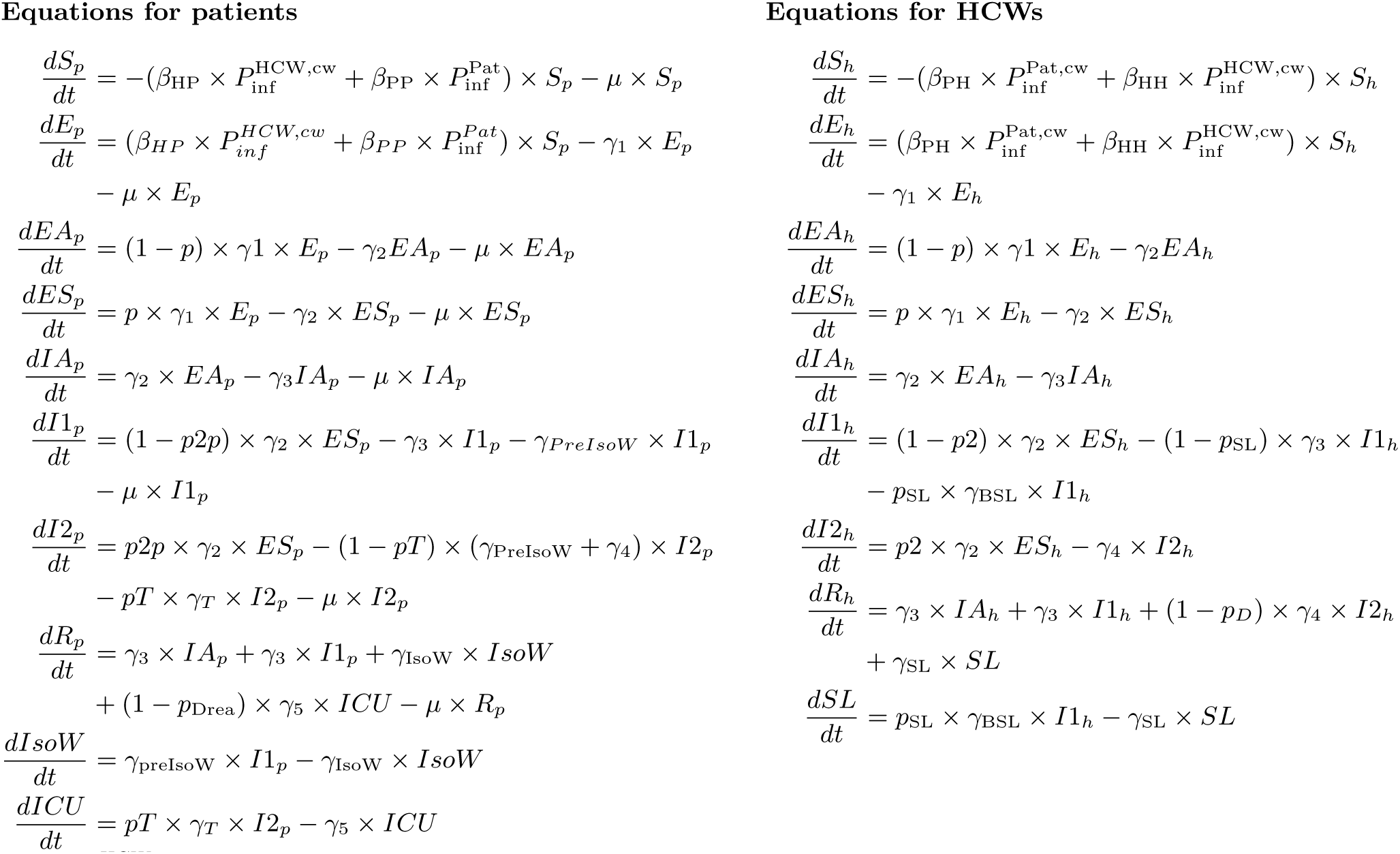

where 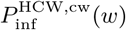 denotes the proportion of infected HCWs in contact with patients belonging to a given ward *w*. We denote the total number of wards by *W* ∈ ℕ. We first compute the proportion of infected HCWs in each ward as follows for *w* = 1, …, *W* :

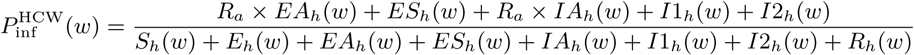

where *R*_*a*_ is the relative risk of transmission of individuals in the asymptomatic pathway relative to individuals in the symptomatic pathway. For simplicity, we omit the ward notation for compartments going forward. The proportion of infected HCWs in contact with patients belonging to a given ward *w* is therefore computed as follows:

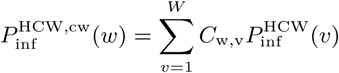

where *C* ∈ ℳ_*W × W*_ (ℝ _≥0_) is a contact matrix computed based on the connections between all *W* wards which are determined by the sharing of HCWs (Fig. S4d). Each entry corresponds to the proportion of total working hours spent by HCWs from the row ward in the column ward. For instance, HCWs of ward *H2* typically spend 33% of their working time on average in ward *H1*.

The proportion of infected patients in a given ward *w* is computed as follows:

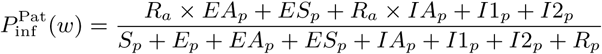

We recall that susceptible HCWs can be infected upon contact with contagious patients and HCWs in all wards they work in. The proportion of infected patients with which HCWs from a given ward *w* are in contact with is therefore computed as follows:

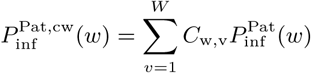

The differential equations which govern the deterministic version of the model with a screening area are given below.

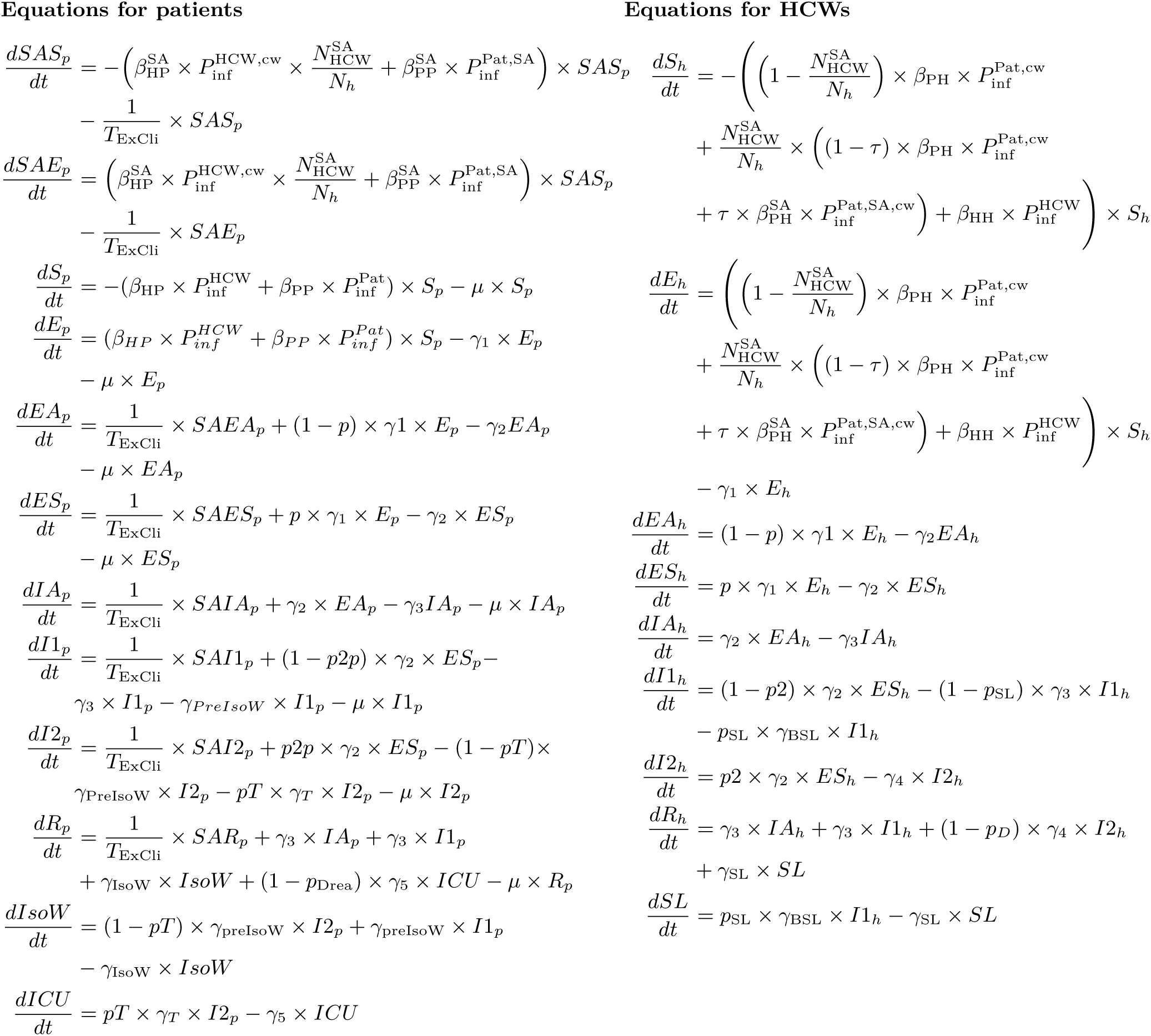

where 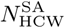 is the total number of HCWs taking care of patients in the screening area of a given ward and *N*_*h*_ is the total number of HCWs working in the ward. 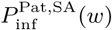 is the number of patients infected in the screening area of a given ward *w* and is calculated as follows:

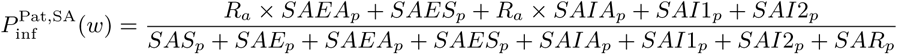

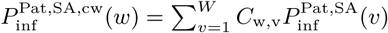 is the proportion of infected patients in screening areas where HCWs of a given ward *w* are working in. *τ* represents the proportion of patients in the screening area of a ward.

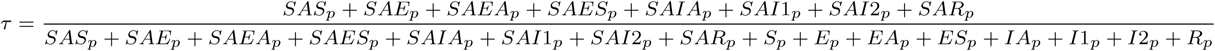

### Statistical inference

#### Transmission rates

Transmission rates *β*_PP_, *β*_HH_, *β*_HP_ and *β*_PH_ were estimated based on an outbreak that occurred in ward A2. The model defined with four transmission rates was not identifiable in simulation settings, resulting in poor recovery of known parameter values. Instead, we defined each transmission rate as a certain constant times *β* and then proceeded with the estimation of the single rate *β*. Multiple combinations were tested and the set that subsequently maximized the likelihood was defined as follows. Taking into account HCW working hours which we assumed to be on average 25 % of the time, and assuming a similar contact rate in-between and between patients and HCWs, we came up with the following ratios: *β*_PP_ = *β*_PH_ = *β, β*_HH_ = 0.25 × *β* and *β*_HP_ = 0.7 × *β*.

#### Likelihood based inference with *pomp*

We implemented a within-ward model parametrized and initialized to match patient and HCW populations’ structure in ward A2 during that period. The likelihood-based inference framework for the stochastic model was provided by iterated filtering methods (1) which were readily implemented in the R package *pomp* (2). The *pomp* object was constructed by specifying the model observations as cumulative symptomatic infections in the patient and HCW populations, the spread of infection based on a Gillespie algorithm using *rprocess* to define the observation process, the measurement model defined as Poisson distributed with parameter equal to the cumulative infections for patients and HCWs using *rmeasure*. Finally, *dmeasure* which is an evaluator of the measurement model was defined analogously to *rmeasure*. Following each time step, the observation process provides a likelihood of observing the data given the internal state of the system. The total likelihood was then computed as the product of likelihood values at each time step. Unknown parameters were estimated by maximum likelihood, using particle filtering to compute a robust estimate of the likelihood and iterated filtering to maximize it over unknown parameters. Particle filtering uses a set of particles to represent the posterior distribution of the stochastic process given noisy observations. Each particle has a likelihood weight assigned to it that represents the probability of that particle being sampled from the probability density function. Resampling is performed at each observation to produce copies of the few particles with the highest weights. The method was implemented using the *pfilter* function and outputs a stochastic estimate of the likelihood. The particle filter was applied a few times to obtain an estimate of the variability, typically ten times with a high number of particles, 10^6^ in our case. The idea behind the iterated particle filtering algorithm is to apply a particle filter to the model in which the parameter vector for each particle is subjected to random perturbations at each observation. In doing so, the parameter vector is made to follow a random walk. As the iterations progress, the intensity of the perturbations is decreased according to a cooling schedule. For instance, we set the cooling fraction such that after 50 iterations, the perturbations are reduced to half their original magnitudes. The iterated filtering algorithm was implemented in the function *mif2* of *pomp*.

#### Preliminary validation of the procedure using synthetic data

The model and statistical inference framework were first validated on synthetic data generated from a range of known parameter values which were then estimated. For each set of parameter values, we generated 50 independent stochastic datasets using the *rmeasure* functionality of *pomp* with each one of them consisting of at least one case in both patient and HCW populations over the observation period. Parameters were then recovered using the iterated filtering algorithm in function *mif2* with the number of particles and number of iterations set at values of 1500 and 500 respectively. We estimated parameter *β* in multiple scenarios with 1 or 2 non-contagious incubating patients and HCWs as index cases and different times of introduction of the index cases (day 1, day 5 and day 10). In all scenarios, estimates for *β* were adequately close to the true value on average as shown in Fig. S1.

#### Parameter estimation on real outbreak data

Following validation of our estimation framework on synthetic data, we analyzed the real outbreak data of ward A2. We estimated the transmission rate *β*, initial number of non-contagious incubating patients *Ep*_0_ and initial number of non-contagious incubating HCWs *Eh*_0_ by building likelihood profiles for each parameter. In doing so, we constructed 95 % confidence intervals for each parameter using the chi-square approximation to the likelihood ratio statistic. For instance, for a given parameter, say *β*, we computed the likelihood using the particle filtering algorithm for a range of fix values of *β* while estimating *Ep*_0_ and *Eh*_0_ in every step. A 95 % confidence interval was then determined as all values above the highest likelihood minus half of the 95 % quantile of the *χ*^2^-square distribution with two degrees of freedom. We repeated this procedure for times of introduction of the index cases (*t*_init_) ranging from 1 to 20 days before the first observed symptomatic patient. The set of parameter values that maximized the likelihood was then retained as estimates. The profile likelihoods coupled with 95 % confidence intervals for best set of parameter estimates are shown in Fig. S2. The best fit suggested an index case 12 days prior to the detection of the first symptomatic patient. The set of parameter estimates that maximized the likelihood were then fed to the transmission model to generate 1000 datasets. We then plotted the mean of those simulations to compare with the observed data for patients in Fig. S3b and observed data for HCWs in Fig. S3b.

**Table S1.** Description of model parameters.

|  | Symbol | Parameter | Value |
| --- | --- | --- | --- |
| Transmission rates | $\beta_{PP}$ | effective transmission rate between patients | 0.4 (Estimated) |
| | $\beta_{HH}$ | effective transmission rate between HCWs | 0.1 (Estimated) |
| | $\beta_{HP}$ | effective transmission rate between contagious HCWs and susceptible patients | 0.28 (Estimated) |
| | $\beta_{PH}$ | effective transmission rate between contagious patients and susceptible HCWs | 0.4 (Estimated) |
| | $\beta_{PP}^{SA}$ | effective transmission rate between patients in screening area | 0.4 (Estimated) |
| | $\beta_{HH}^{SA}$ | effective transmission rates between HCWs in screening area | 0.1 (Estimated) |
| | $\beta_{HP}^{SA}$ | effective transmission rates between contagious HCWs and susceptible patients in screening area | 0.28 (Estimated) |
| | $\beta_{PH}^{SA}$ | effective transmission rates between contagious patients and susceptible HCWs in screening area | 0.4 (Estimated) |
| | $R_a$ | relative risk of secondary attack rate of people with asymptomatic infections | 0.35 (3) |
| Durations (in days) | $1/\gamma_1$ | duration of non-contagious incubation period | 5 (4) |
| | $1/\gamma_2$ | duration of contagious incubation period | 2 (4) |
| | $1/\gamma_3$ | duration of contagious period when asymptomatic and non-severe symptoms | 7 (4) |
| | $1/\gamma_4$ | duration of contagious period when severe symptoms | 8 (Assumption) |
| | $1/\gamma_5$ | duration of stay in intensive care before recovery or death | 10 (Hospital data) |
| | $1/\gamma_T$ | duration of contagious period before transfer in intensive care when severe symptoms | 2 (Hospital data) |
| | $1/\gamma_{BSL}$ | duration before sick leave when mild symptoms | 1 (Hospital data) |
| | $1/\gamma_{SL}$ | duration of sick leave | 14 (Hospital data) |
| | $1/\gamma_{preIsoW}$ | duration between first symptoms and transfer in a specific ward designated to isolate symptomatic patients | 1 (Hospital data) |
| | $1/\gamma_{IsoW}$ | duration of stay in a specific ward designated to isolate symptomatic patients | 11 (Hospital data) |
| | $T_{ExCli}$ | duration of stay in screening area for clinical examination | $1/24$ (Hospital data) |
| | $T_{Test}$ | duration of stay in screening area for RT-PCR testing | $1/24$ (Hospital data) |
| Probabilities | $p$ | probability of symptoms | 0.7 (3) |
| | $p_2$ | probability of severity if symptoms for HCW | 0.2 (Assumption) |
| | $p_{2p}$ | probability of severity if symptoms for patient | 0.5 (Assumption) |
| | $p_{Drea}$ | probability of death in ICU following severe symptoms for patient | 0.5 (Assumption) |
| | $p_D$ | probability of death following severe symptoms for HCWs | 0.1 (Assumption) |
| | $p_T$ | probability of being transferred following severe symptoms | 0.2 (Assumption) |
| | $p_{SL}$ | probability that HCWS take sick leave following symptoms | 0.8 (Hospital data) |
| | $Prev_{Com}$ | prevalence in general population | 0.1 % - 3 % |
| Testing | $Sens_{Inf}$ | sensitivity of RT-PCR test when testing contagious individuals | 0.8 (5) |
| | $Sens_{NInf}$ | sensitivity of RT-PCR tests when testing non-contagious individuals | 0.3 (5) |
| | $Spec$ | specificity of RT-PCR tests | 1 (5) |
| | $\mu$ | probability of discharge or death | 0.00749 - 0.0547 (Hospital data) |

**Table S2.** Description of each hospital ward including number of HCWs shared between wards.

| Ward | Number of patients | Number of assigned HCWs | Number of shared HCWs | Average stay of patients (in days) | Degree (number of connected wards) |
| --- | --- | --- | --- | --- | --- |
| A1 | 20 | 18 |  | 18.3 | 6 |
| A2 | 20 | 16 | 89 | 133 | 6 |
| A3 | 25 | 25 |  | 27.4 | 5 |
| A4 | 20 | 19 |  | 50.8 | 5 |
| A5 | 25 | 21 | 47 | 21.9 | 6 |
| A6 | 20 | 18 |  | 21.6 | 5 |
| B1 | 20 | 22 |  | 82.7 | 6 |
| B2 | 23 | 20 | 31 | 40.9 | 2 |
| E1 | 25 | 20 |  | 35.9 | 2 |
| C1 | 20 | 16 |  | 53.5 | 2 |
| D1 | 20 | 18 | 38 | 20.3 | 2 |
| D2 | 20 | 16 |  | 19.1 | 2 |
| C2 | 19 | 18 |  | 27.8 | 4 |
| C3 | 20 | 18 |  | 18.8 | 4 |
| E2 | 28 | 22 | 56 | 41.6 | 7 |
| F1 | 24 | 22 |  | 64.6 | 7 |
| F2 | 24 | 24 |  | 147 | 7 |
| B3 | 22 | 18 |  | 18.5 | 6 |
| B4 | 19 | 16 | 52 | 30.7 | 6 |
| C4 | 19 | 13 |  | 19.8 | 6 |
| G1 | 18 | 17 | 39 | 32.7 | 3 |
| G2 | 19 | 17 |  | 25.7 | 1 |
| H1 | 25 | 19 | 38 | 26.3 | 2 |
| H2 | 24 | 21 |  | 24.5 | 2 |

**Fig. S1.**
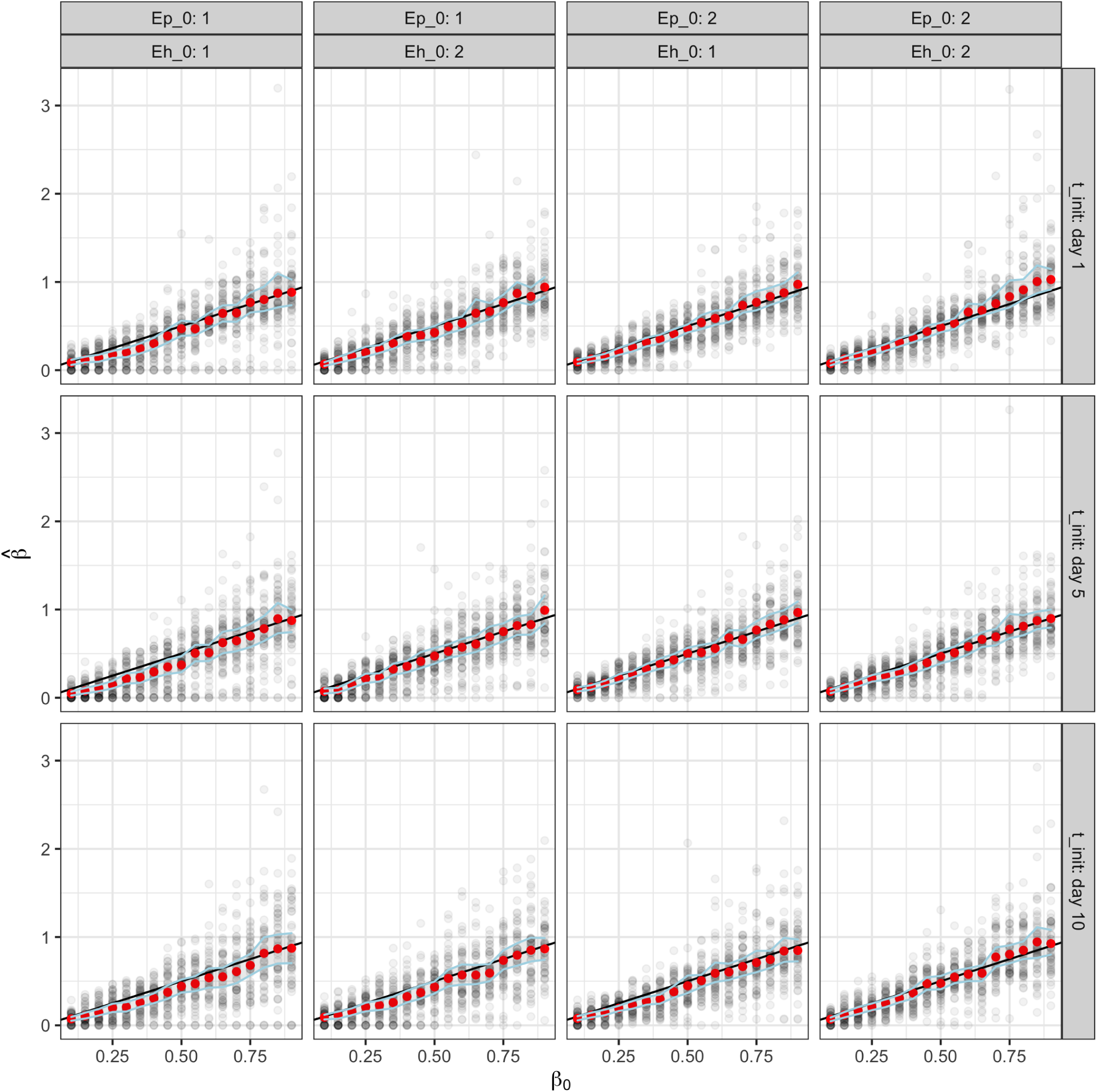
Mean estimates of *β* in multiple scenarios. Mean estimates of *β* in red with 95 % confidence intervals based on 50 simulations in each instance. The number of patients 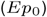 and HCWs 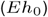 in non-contagious incubation on the first day are fixed at values indicated in the column facet labels. Row facet labels indicate the date of introduction of the index cases.

**Fig. S2.**
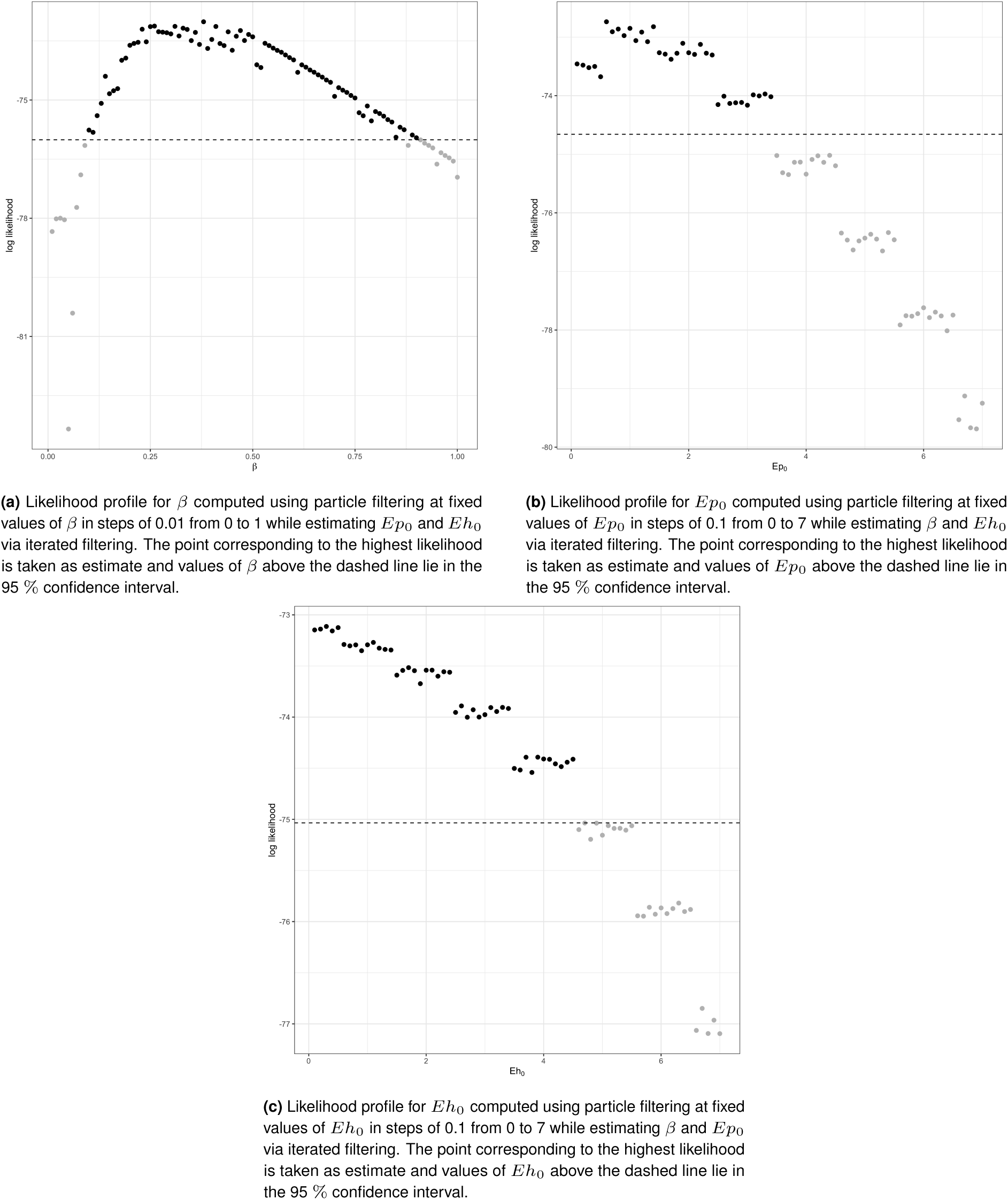
Likelihood profile for parameters *β*, 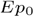 and 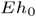 computed on real outbreak data.

**Fig. S3.**
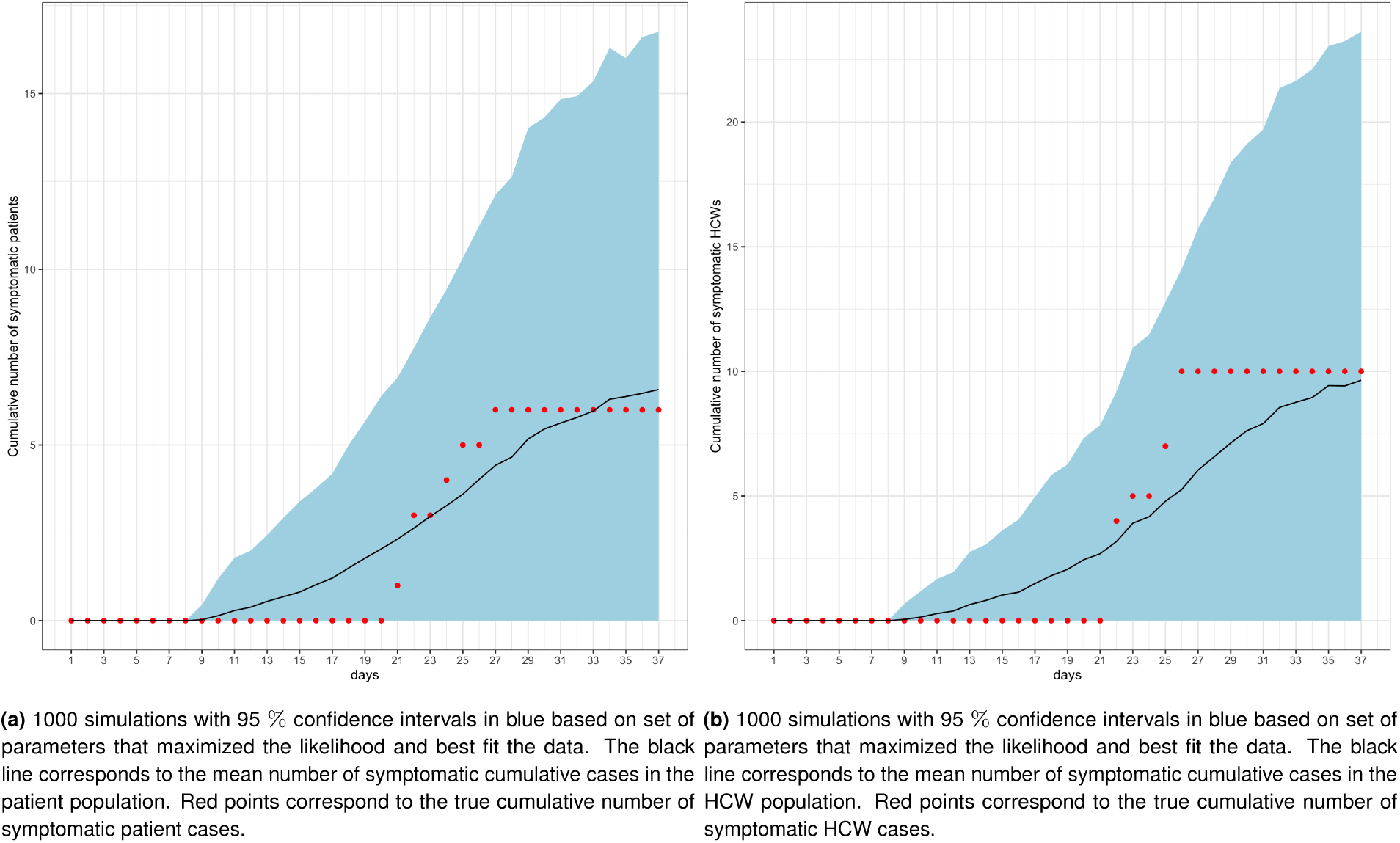
Simulating patient and HCW data using best set of parameter estimates. Cumulative incidence over time (a) in patients and (b) in HCWs

**Fig. S4.**
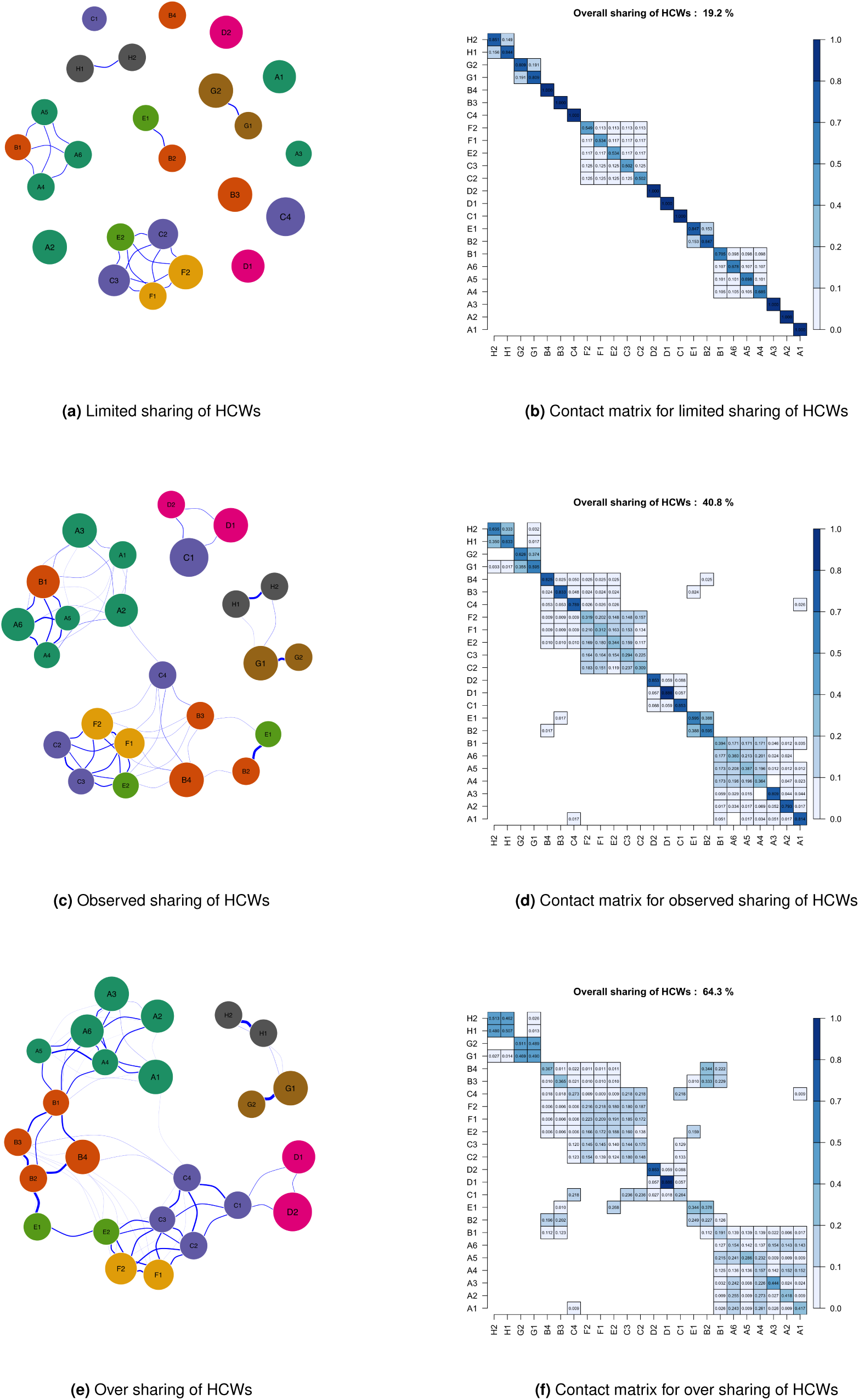
Wards located in the same building are color coded similarly. From top to bottom, network representation of the sharing structure on the left with respective contact matrix on the right.

## Supplementary simulation results

**Fig. S5.**
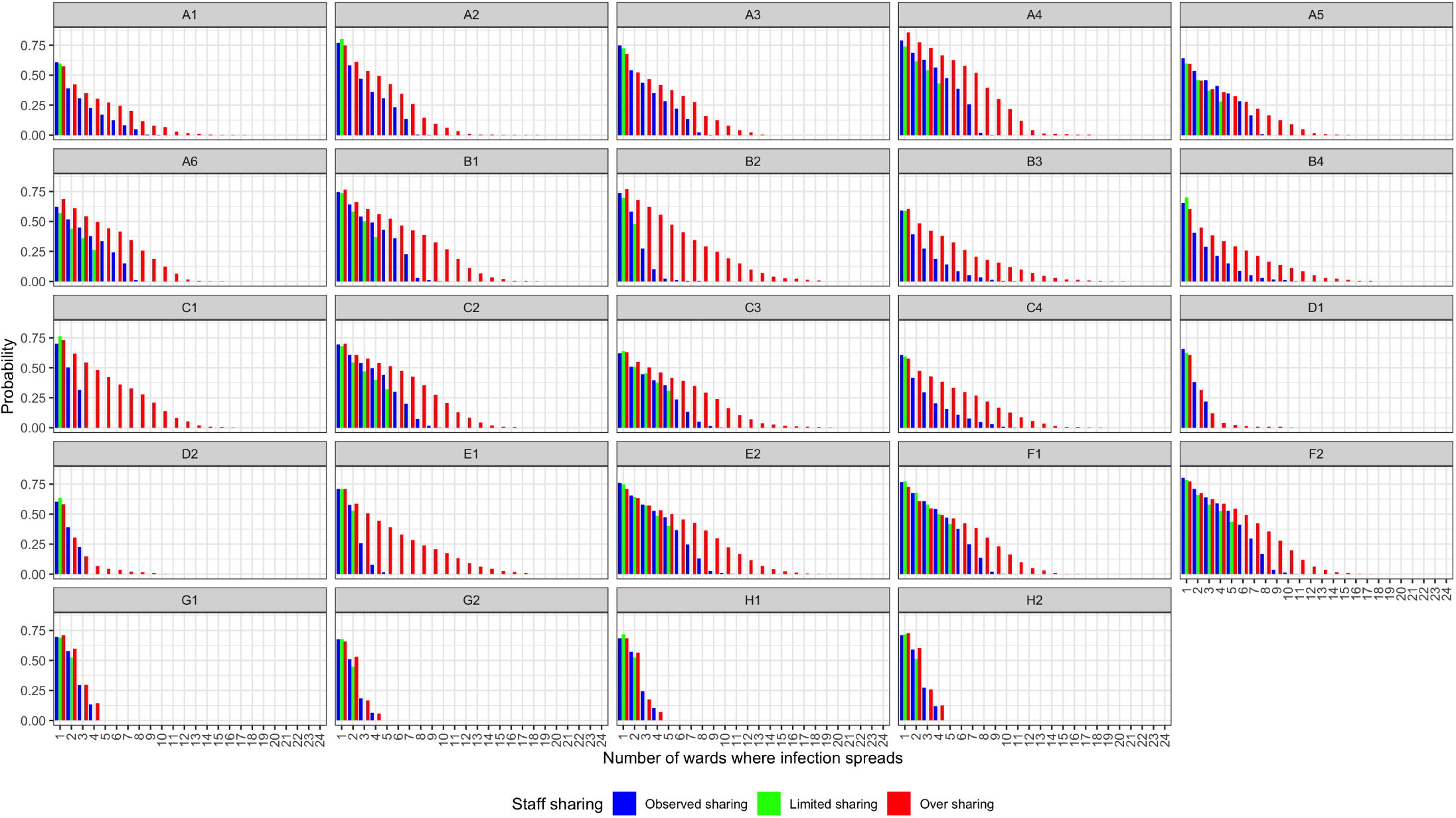
Impact of altering HCWs sharing matrix on the number of wards affected after the introduction of an index case. Probability of spread (y-axis) in number of wards ranging from 1 to 24 (x-axis) for the three staff organization levels based on 500 simulations over a period of 40 days. Facet labels correspond to index wards. Context of simulation: No possible contamination from the community. Introduction of a non-contagious incubating patient in each ward.

**Fig. S6.**
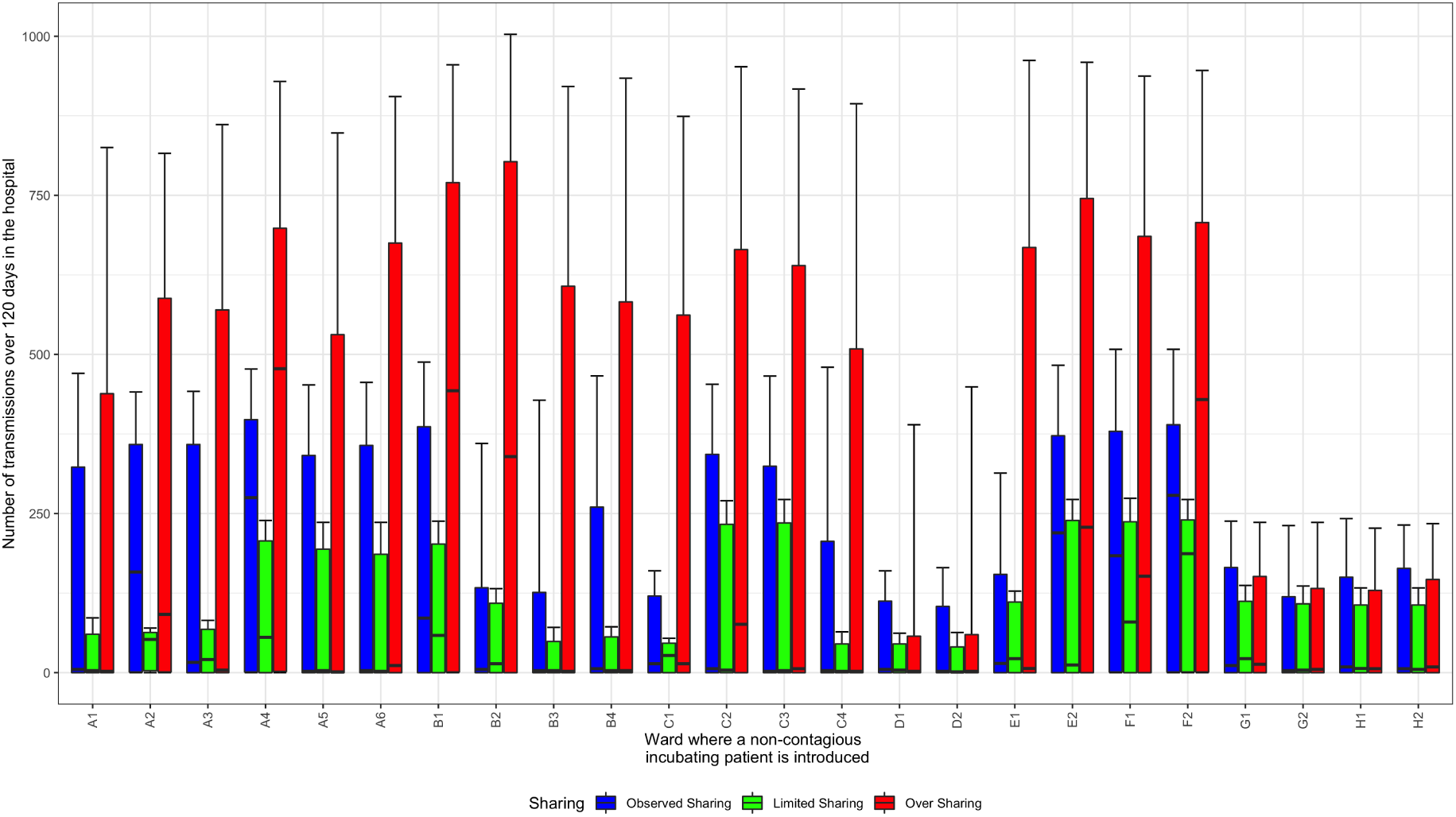
Risk of dissemination at the hospital level depending on the ward of introduction. Number of transmissions following the introduction of an index case in each ward for three staff organization levels based on 500 simulations over a period of 120 days. No contamination from the community considered.

**Table S3.**
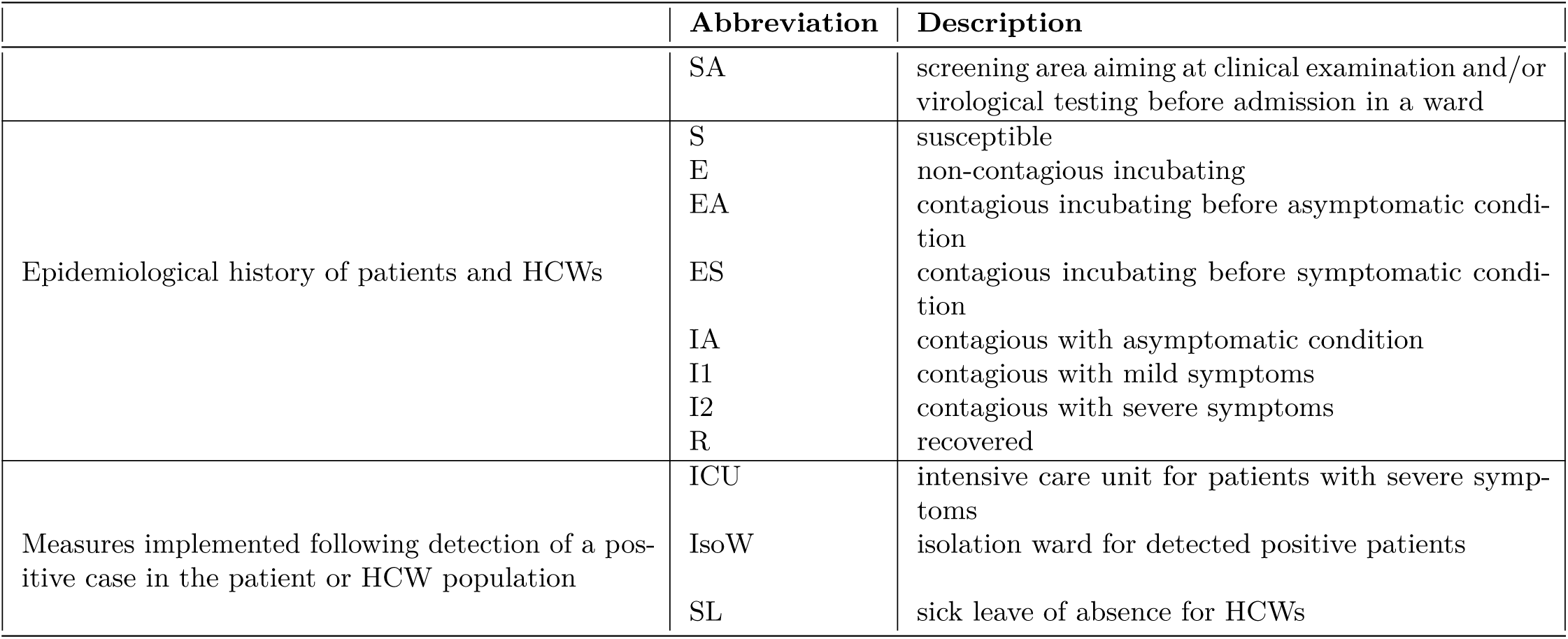
Description of model compartments.

